# Automated Video-Based Analysis of Surgical Meta-competencies Using Computer Vision

**DOI:** 10.1101/2025.11.24.25340912

**Authors:** Josiah Aklilu, Joshua A. Villarreal, Chloe K. Nobuhara, Charlotte Egeland, Xiaohan Wang, Elaine Sui, Alan Brown, Matthew Leipzig, Reid Dale, Anita Rau, Alfred Song, Shelly Goel, Eric Sorenson, Vanessa Palter, Roger Bohn, Teodor Grantcharov, Jeffrey K. Jopling, Serena Yeung-Levy

## Abstract

**Background:** Traditional surgical training relies on an apprenticeship model, which is subjective and threatened by human bias. Performance metric scales attempt to offer more objective feedback by providing a structured grading rubric, but these scores are still ultimately subjective. Leveraging computer vision and artificial intelligence to assess surgical performance has the potential to shift subjective measurements into automated and objective feedback for trainees.

**Materials and Methods:** This retrospective, multi-institutional study analyzed 319 laparoscopic cholecystectomy videos from IRB-approved deidentified datasets and segmented the videos into 2862 clips. Using an internally validated video-based assessment rubric, we annotated video clips across five metacompetency domains: tissue handling, psychomotor skills, efficiency, dissection quality, and exposure quality. Short video segments (*<*90s) were rated on a 5-point scale by expert raters. We trained a deep learning model (DINOv2) to classify composite high (4–5) vs low (1–3) metacompetency scores, representing yes/no binary feedback, a realistic comparison to the operating room. Model performance was evaluated via area under the receiver operating characteristic curve.

**Results:** Among 2862 LC video clips, model performance was highest for dissection quality during the exposing gallbladder step (AUROC 91.5%, 95% confidence interval [CI], 84.5-96.5). Moderate performance was observed for efficiency (AUROC 72.6%, 95% CI 59.9-83.2) and exposure quality (AUROC 68.7%, 95% CI 55.2-81.8). Dissection and exposure quality scores during hepatocystic triangle dissection yielded AUROCs of 63.8% (95% CI 56.9-71.3) and 66.0% (95% CI 53.1-76.7), respectively.

**Conclusion:** We demonstrate the feasibility of a purely vision-based deep learning model to grade surgical skill based on metacompetencies with excellent performance during simple steps. This technique represents an advance over prior whole-video approaches that rely heavily on tool- tracking and kinematic data, and may lead to greater model performance by using binary feedback on increasingly specific step segmentation.

## Introduction

The foundation of surgical education is built on a master-apprentice relationship between faculty and trainees (Camison et al. 2022). This tried and true paradigm, while effective, has the potential to introduce subjective measurements and bias into surgical training (Gerull et al. 2019; Vogt et al. 2003). Human bias in training can be favorable or unfavorable, and master surgeons can be influenced by their own surgical preferences in technique or the nature of the apprentice (Vogt et al. 2003). To combat the subjective nature of training, several objective rubric-style measurements have been introduced in the past few decades. Objective Structured Assessment of Technical Skills (OSATS) and Entrustable Professional Activities (EPAs) represent two recent shifts towards measuring surgical competency in a more objective fashion (Hiemstra et al. 2011; Hatala et al. 2015; Brasel et al. 2023). These grading scales can now provide numerical feedback, but are still influenced by a rater’s human bias (Sherbino and Norman 2017).

Artificial intelligence (AI) presents a unique opportunity to leverage the unbiased nature of computer programming to create truly objective and automated performance metrics (Ryder et al. 2024; Igaki et al. 2023). Furthermore, AI is scalable; while the master-apprentice model requires oneto-one supervision, AI has the potential to automate assessment with immediate feedback to multiple individuals simultaneously. Previously, other groups have shown that AI can provide automated, objective feedback of surgical skills with comparable discrimination to a human reviewer (Kiyasseh et al. 2023; Lavanchy et al. 2021). However, prior work has been limited in its performance by analyzing and scoring a surgical video based on a procedure in its entirety and relying heavily on tool-tracking and kinematic data (Kiyasseh et al. 2023). We hypothesized that by segmenting a procedure into steps, we would be able to achieve higher agreement in human and model performance. Master surgeon raters would have higher levels of disagreement in more complex steps and higher levels of agreement (and therefore, better model performance) in simpler steps.

We sought to address this question by taking a well-characterized cohort of 319 laparoscopic cholecystectomy (LC) videos and dividing them into 2862 video clips stratified by procedure step. We then trained a deep learning model to classify technical performance across five metacompetency (MC) domains: tissue handling, psychomotor skill, efficiency, dissection quality, and exposure quality (Igaki et al. 2022). With these divisions in mind, the model was trained to judge each 45-70 second clip into a binary classifier of high or low performance, with high being an MC score of 4-5 and low being an MC score of 1-3, and we validated its performance with human ratings. This is, to our knowledge, the first study to show purely vision-based performance metrics of surgical metacompetencies in laparoscopic cholecystectomy.

## Materials and Methods

### Video Dataset Collection and Annotations

This multi-institutional retrospective study included a dataset of 319 LC composed of de-identified publicly available and Institutional Review Board (IRB) approved intraoperative videos from two United States academic medical centers. We used an established surgical video annotation framework for video annotations and internally validated a video-based assessment (VBA) rubric for LC using validated universal surgical MC (Meireles et al. 2021). A hierarchical task analysis for LC was conducted to define the intracorporeal steps for MC review. Ratings covered five skill domains, namely tissue handling, psychomotor skills, efficiency, dissection quality, and exposure quality. Meta-competency ratings were assigned on a 5-point Likert scale by a team of four expert raters (Table 1).

**Table 1:** Characterization of video clip dataset. We report the total number of video clips from our mutli-source dataset and the number of unique procedures the clips were extracted from.

| Labeled datasets | No. video clips | No. unique procedures |
| --- | --- | --- |
| Proprietary dataset 1 | 759 | 139 |
| Proprietary dataset 2 | 620 | 77 |
| Cholec80 | 649 | 80 |
| Heichole | 210 | 23 |
| **Total clips** | **2862** |  |

**Table 2:**
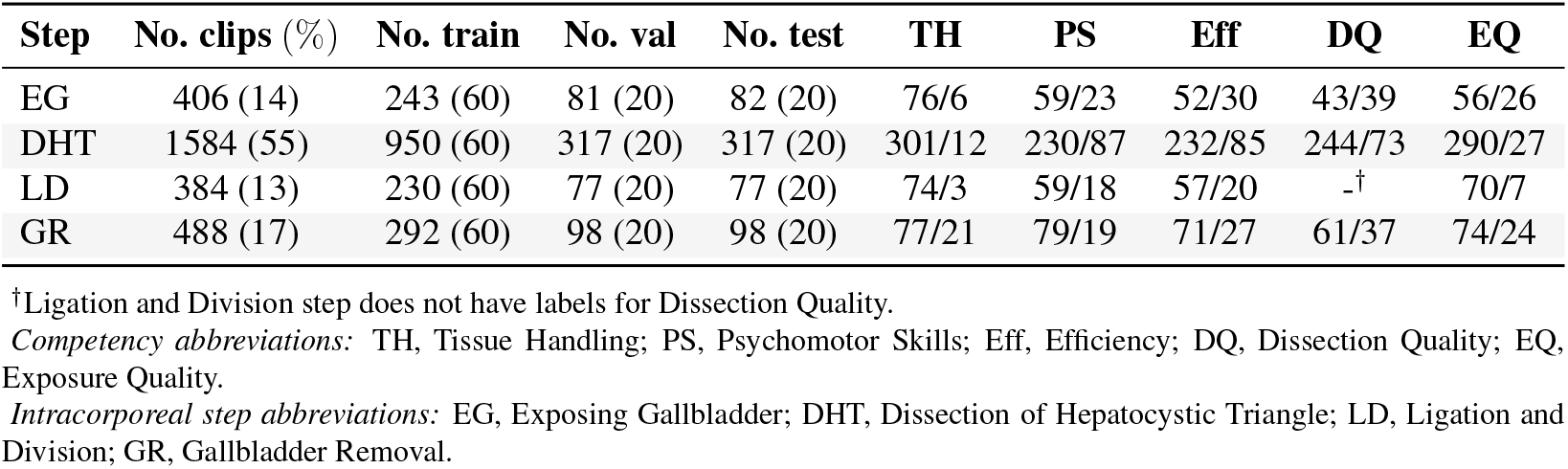
Surgical meta-competency annotations. Total counts of video clips from each step of laparoscopic cholecystectomy, and training and test splits for model development. We use a 60:20:20 split for all competency models. Counts in the meta-competency columns represent the number of *high*vs *low*-skill samples (respectively) included per procedural step on the test set.

| Step | No. clips (%) | No. train | No. val | No. test | TH | PS | Eff | DQ | EQ |
| --- | --- | --- | --- | --- | --- | --- | --- | --- | --- |
| EG | 406 (14) | 243 (60) | 81 (20) | 82 (20) | 76/6 | 59/23 | 52/30 | 43/39 | 56/26 |
| DHT | 1584 (55) | 950 (60) | 317 (20) | 317 (20) | 301/12 | 230/87 | 232/85 | 244/73 | 290/27 |
| LD | 384 (13) | 230 (60) | 77 (20) | 77 (20) | 74/3 | 59/18 | 57/20 | -<sup>†</sup> | 70/7 |
| GR | 488 (17) | 292 (60) | 98 (20) | 98 (20) | 77/21 | 79/19 | 71/27 | 61/37 | 74/24 |

<sup>†</sup>Ligation and Division step does not have labels for Dissection Quality.
*Competency abbreviations:* TH, Tissue Handling; PS, Psychomotor Skills; Eff, Efficiency; DQ, Dissection Quality; EQ, Exposure Quality.
*Intracorporeal step abbreviations:* EG, Exposing Gallbladder; DHT, Dissection of Hepatocystic Triangle; LD, Ligation and Division; GR, Gallbladder Removal.

### Model Architecture and Training

Our model architecture consisted of a DINOv2 (Oquab et al. 2024), a pretrained vision transformer backbone (ViT-B) and a multi-stage temporal convolutional network (TCN) with 4 TCN layers (Lea et al. 2016). DINOv2 is an advanced artificial intelligence (AI) system for image analysis. It learns to identify important visual features and patterns by analyzing millions of general, nonmedical images on its own, without requiring manual labeling by human experts. We leveraged DINOv2, which was trained on a corpus of 142 million web-crawled images, to produce highly-descriptive features of each video frame in the surgical clips. The TCN is another type of computer vision-based AI system that learns temporal relationships. We used TCN to process the DINOv2 features extracted from each frame, allowing the model to learn temporal relationships between video frame features throughout a single clip. Thus, each meta-competency model comprised of a TCN over video frame features extracted by DINOv2 for each clip.

To train these models, we used Low-Rank Adaptation (LoRA) [Hu et al. 2021] fine-tuning for the DINOv2 vision backbone and full parameter fine-tuning of the TCN head. LoRA is a computationally efficient method to specialize large AI models for new tasks. Instead of retraining the entire model, LoRA freezes the original model’s knowledge and trains only a small set of new parameters. This approach significantly reduces the computational resources required and, crucially, prevents the loss of the original model’s powerful foundational knowledge when adapting it to a specialized dataset. In our study, we leveraged the powerful, general-purpose representations from the DINOv2 model while specializing it for our surgical video dataset, which has a distinct visual domain. Also for training, we used the AdamW optimizer for adaptively updating our model parameters with each training step. This optimizer allows the model to adjust individual parameters at different rates to adjust its outputs for each batch of video clips it sees during training. We use a learning rate of 1*e*^−6^ for the backbone and 3*e*^−5^ for the TCN head (Figure 1). We also implemented weight decay of 0.01 for the backbone and 0.05 for the TCN head, along with dropout rate of 0.3 for regularization. These two techniques prevent the model from memorizing training videos clips and help with generalization to new videos. We trained our models for 20 epochs on our training datasets. The models were then trained to differentiate between a binarized composite of low (1-3) and high (4-5) levels of operative performance according to MC ratings.

**Figure 1:**
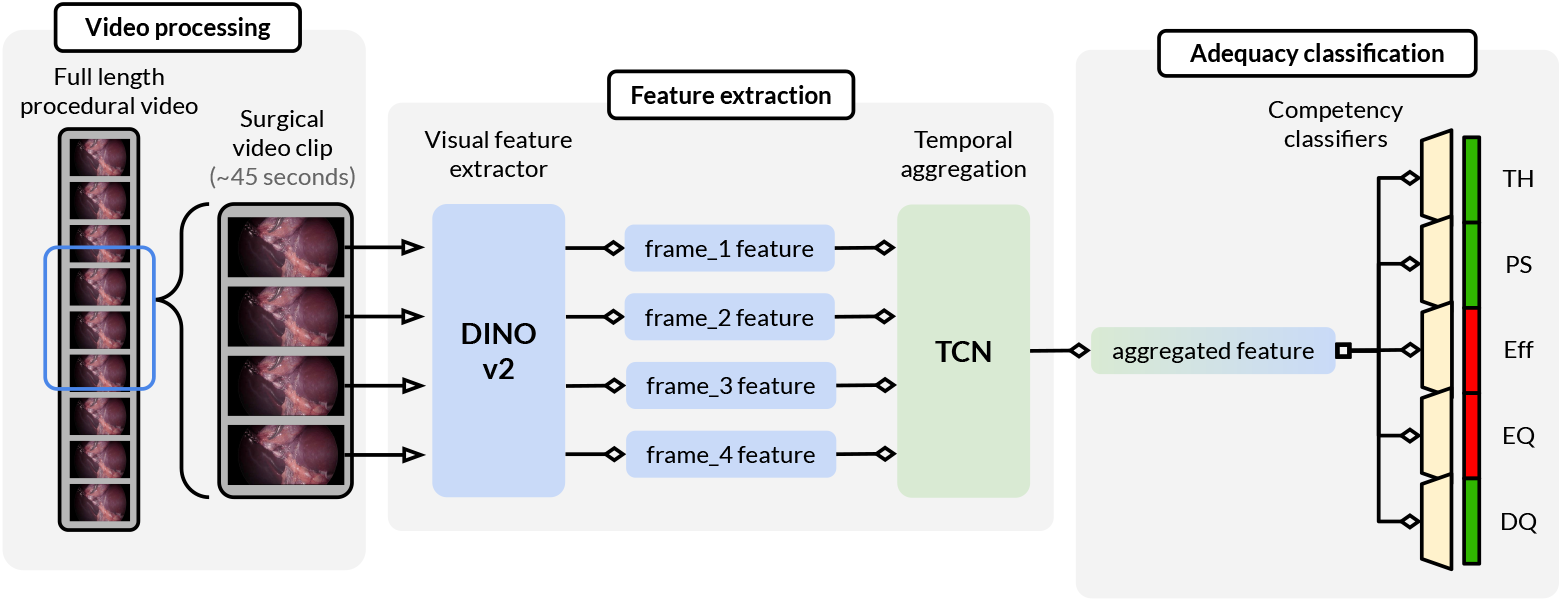
Computer Vision Model Architecture. Design of the computer vision model used to classify adequacy in each surgical meta-competency.

## Results

Our dataset used a total of 319 intraoperative, deidentified videos of LC from two United States academic medical centers (Proprietary dataset 1, Proprietary dataset 2) and international surgical collaborators (Cholec80, Heichole) [Table 1]. The initial stage of clip generation involved first segmenting each video into four steps of the LC procedure: (1) Expose gallbladder, (2) Dissection of hepatocystic triangle, (3) Ligation and division, (4) Gallbladder removal. Segments longer than 360 seconds were divided into four quartiles, with one 60–90 second clip drawn from each. A maximum clip duration of 90-seconds was chosen as the high end value as an interval representing the practical timeframe for video annotators to rate a single surgical maneuver. This clip generation procedure created 2891 clips, of which 2862 were usable for annotations from human reviewers (e.g. 29 clips were excluded for lack of active operating, poor image quality, or extracorporeal footage) [Table 1].

All 2862 clips received annotations from human reviewers on the five metacompetency (MC) domains: tissue handling, psychomotor skill, efficiency, dissection quality, and exposure quality. A subset of dataset was labeled by multiple reviewers, with a planned overlap in assignments to allow for the measurement of inter-rater reliability. We reported model discrimination performance with area under the receiver operating characteristic curve (AUROC) in our testing data. We employed AUROC as our primary evaluation metric because it offers a robust measure of the classifier’s performance across a range of decision thresholds. Additionally, we used 1000 bootstrap samples to compute all 95% confidence intervals in our analysis.

Our model achieved its highest performance for the exposing gallbladder (EG) step, achieving an AUROC of 91.5% (95% confidence interval [CI], 84.5-96.5) for discriminating dissection quality (Figure 2). Additionally, for the EG step, the model demonstrated modest performance for efficiency (AUROC 72.6%, 95% CI, 59.9-83.2) and exposure equality (AUROC 68.7%, 95% CI, 55.2-81.8). For the dissection of hepatocystic triangle (HCT) step where identification of the critical view of safety is achieved, our model demonstrated an AUROC of 66.0% (95% CI, 53.1-76.7) in classifying exposure quality and 63.8% (95% CI, 56.9-71.3) for dissection quality.

**Figure 2:**
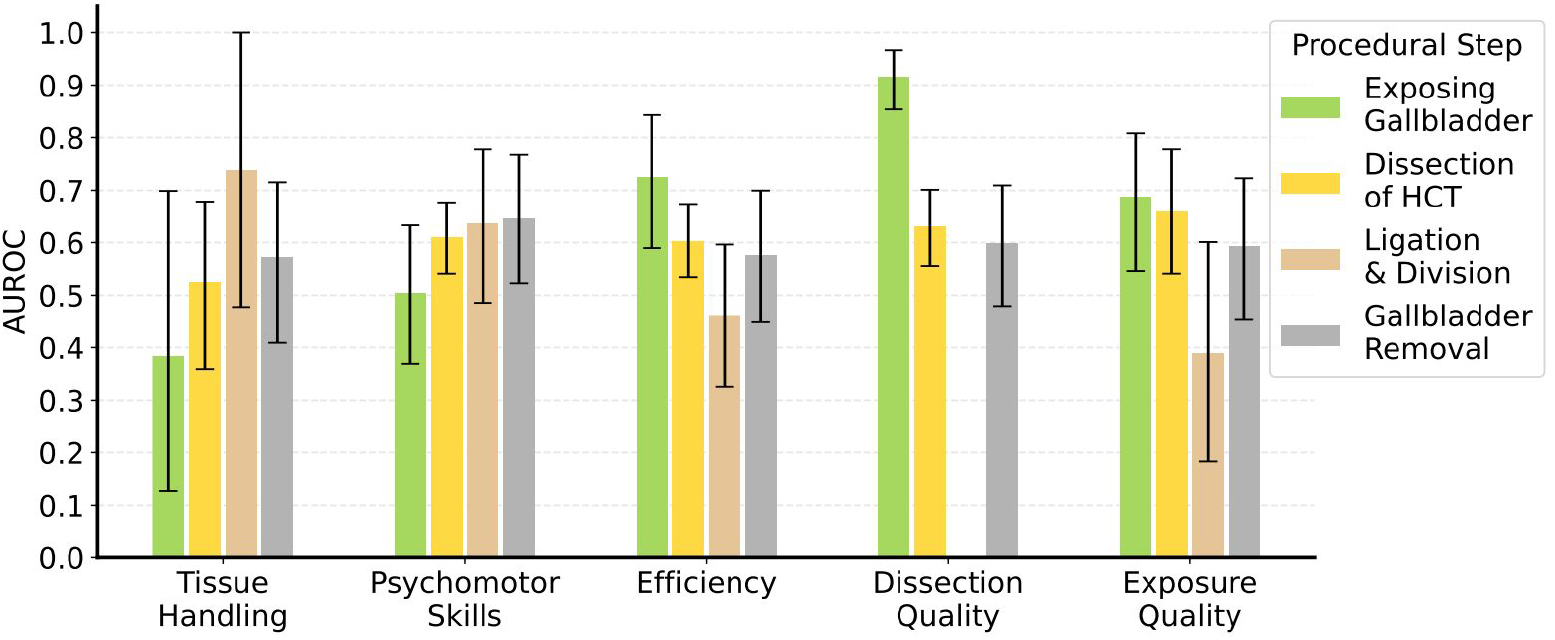
Model Discrimination Metrics for Meta-competency Predictions Across Cholecystectomy Steps. We plot the performance for each of our meta-competency models per intracoproreal step. Note that the Ligation & Division step does not have ratings for dissection quality since no dissection occurs during this step. Error bars indicate 95% confidence intervals. *Abbreviations:* HCT, hepatocystic triangle.

## Discussion

Surgical pedagogy has traditionally relied on a one-to-one training and expert opinion, which is subjective, threatened by human biases, and limited in its scalability. In the age of artificial intelligence (AI), there is a unique opportunity to capitalize on the unbiased and calculated nature of machine learning to transfer performance metrics from what is currently a subjective art into an objective science. Here, we tested the feasibility of using a pure vision-only approach to classify laparoscopic cholecystectomy (LC) into high versus low skill, by segmenting the LC procedure into key steps, and found excellent model agreement in simpler steps. More complex steps may need further subdivision to achieve this level of agreement between AI and human raters.

Currently, AI has been used in other proof-of-concept studies to automate surgical performance metrics. These studies either use kinematics (e.g. physical data from or either motion sensors, force feedback, or three-dimensional coordinates from robotics) or computer vision (e.g. using programmed algorithms to understand videos in real time and extract data) [Kiyasseh et al. 2023; Soangra et al. 2022]. Following data collection, there are several performance metrics where automated analysis can be completed using machine learning. Deep learning, a subset of machine learning, utilizes multi-layered architectures inspired by the human brain (i.e. neural networks) to learn increasingly complex features from high-dimensional video (Pedrett et al. 2023). We used DINOv2, a deep learning algorithm trained on a curated dataset of *>*140 million images and that excels in tasks requiring temporal/spatial understanding to produce descriptive visual features for each surgical video clip (Oquab et al. 2024).

Other studies have primarily focused on used machine learning and a combination of kinematics or tool tracking data to quantify performance metrics (Khalid et al. 2020; Kiyasseh et al. 2023; Lam et al. 2022; Lavanchy et al. 2021). Few studies have used a ordered scale of proficiency to train a model to recognize level of skill based on a metacompetency (MC)-based metrics (Pérez-Escamirosa et al. 2020; Pan et al. 2023). No study to date has then segmented the procedure into clips to then create a binary readout of high skill (MC 4-5) or low skill (MC 1-3). We initially hypothesized that variations in inter-rater reliability across the entire procedure were high because the operator can perform with both high skill and low skill in the same operation, and raters may anchor to one demonstration more than the other. Therefore, by segmenting the procedure into clips, we would achieve better model performance and excellent performance in the simpler steps (e.g. rating the dissection quality during gallbladder exposure).

While our model demonstrated the feasibility of using computer vision to classify surgical skill with excellent accuracy, only one model achieved excellent performance of over 90% for dissection quality of exposing the gallbladder. Other MC metrics in the EG step achieved modest performance of approximately 70%. Notably, and arguably where evaluating performance matters the most, the dissection of the hepatocystic triangle step reached at highest an AUROC of 66% despite this step being the largest clip dataset at n=950. This indicates that human raters still have high levels of disagreement in assessing objective performance metrics at complex and critical steps in the operation. Furthermore, we used our own internal video-based assessment rubric. External validation of video-based assessments in MC domains (e.g. via a modified Delphi process) can promote standardization in AI assessments (Meireles et al. 2021).

Model development in the era of computer vision and surgical performance is limited by variations in human labels and degree of disagreement. Two surgeons watching a 60-minute clip are likely to grade the overall MCs differently due to internal biases and mental anchoring on a high skill or low skill portion of performance during the procedure. The clip generation portion of this study is a novel approach to using computer vision to extract data from surgical videos, and combining this with a binary performance metric of high versus low skill mirrors the yes/no feedback that a trainee is more likely to hear while operating. From a computer vision standpoint, this approach represents an entirely new workflow and paradigm for facilitating human rater disagreement by segmenting longer videos into shorter segments. From a surgical standpoint, this study represents a proof-of-concept approach to using computer vision and surgical video clips to automate objective feedback and assessment for simple steps of an operation.

## Data Availability

All data produced in the present study are available upon reasonable request to the authors.

